# Combining imaging- and gene-based hypoxia biomarkers in cervical cancer improves prediction of chemoradiotherapy failure independent of intratumor heterogeneity

**DOI:** 10.1101/2020.05.28.20115386

**Authors:** Christina S. Fjeldbo, Tord Hompland, Tiril Hillestad, Eva-Katrine Aarnes, Clara-Cecilie Günther, Gunnar B. Kristensen, Eirik Malinen, Heidi Lyng

## Abstract

**Purpose:** Emerging biomarkers from medical imaging or molecular characterization of tumor biopsies open up for combining the two and exploiting their synergy in treatment planning. We compared pretreatment classification of locally advanced cervical cancer patients by two previously validated imaging- and gene-based hypoxia biomarkers, appraised the influence of intratumor heterogeneity, and investigated the benefit of combining them in prediction of chemoradiotherapy failure.

**Experimental Design:** Hypoxic fraction, determined from dynamic contrast enhanced (DCE)-MR images, and an expression signature of 6 hypoxia-responsive genes were used as imaging- and gene-based biomarker, respectively, in 118 patients. Intratumor heterogeneity was assessed by variance analysis. The biomarkers were combined using a dimension reduction procedure.

**Results:** The two biomarkers classified 75% of the patients with the same hypoxia status. Inconsistent classification in some cases was not related to imaging-defined intratumor heterogeneity in hypoxia, and hypoxia status of the slice covering the biopsy region was representative of the whole tumor. Hypoxia assessed by gene expression was independent on tumor cell fraction in the biopsies and showed minor heterogeneity across multiple samples in 9 tumors. Inconsistent classification was therefore rather caused by a difference in the hypoxia phenotype reflected by the biomarkers, providing a rational for combining them into a composite score. This score showed improved prediction of treatment failure (HR:7.3) compared to imaging (HR:3.8) and genes (HR:3.0) and significant prognostic impact in multivariate analysis with clinical variables.

**Conclusion:** Combining our imaging- and gene-based biomarkers enables more precise and informative assessment of hypoxia-related chemoradiotherapy resistance in cervical cancer.

## Introduction

Advances in medical imaging and molecular characterization of tumors have shown promise for identifying treatment-resistant cancer and deciding therapy (1–3). Incorporation of the two methodologies in the clinic may improve treatment-decision, and is an important step towards precision medicine (3,4). Hypoxia is a major adverse feature of solid tumors, leading to metastases and resistance to radiotherapy, chemotherapy, and possibly molecular targeted drugs and immunotherapies (5,6). Promising imaging- and gene-based hypoxia biomarkers have been proposed, including candidates derived from positron emission tomography (PET) and magnetic resonance (MR) images (7), and gene expression signatures recorded in tumor biopsies (8). In particular, PET with the hypoxia tracer F-18-fluoromisonidazole has shown potential for targeted, local radiation dose escalation in head and neck cancer (9). Also, dynamic contrast enhanced (DCE)-MR imaging (MRI) has shown benefit in monitoring effects of the hypoxia-modifying drug sorafenib in cervical cancer (10). Moreover, gene expression signatures with predictive impact in hypoxia-modifying combination therapies of head and neck and bladder cancer have been presented (11,12) and/or are evaluated further in ongoing intervention trials (NCT01950689, NCT01880359, NCT02661152, NCT04275713). These developments constitute an excellent basis for combining imaging and molecular characterization to exploit advantages of each methodology in an extended treatment decision support system that includes hypoxia.

Imaging- and gene-based hypoxia biomarkers provide different information of value for treatment planning. Imaging can non-invasively visualize hypoxia in three dimensions prior to and during therapy, assess intratumor heterogeneity, and monitor therapy responses repeatedly (13). Approaches based on MR and PET can easily be implemented, since these modalities are part of the state-of-the-art diagnostic procedures for many cancer types, including cervical cancer (14). On the other hand, gene expression signatures capture the transcriptional state of cells and can inform about hypoxia-related resistance mechanisms at play in individual tumors (15). This is of utmost importance for the choice of hypoxia-targeting drug among a large number of existing and upcoming agents for combination therapies (16). A major obstacle is, however, that the information provided in a biopsy may be biased by the cellular composition of the sample and intratumor heterogeneity in hypoxia (17). To exploit the potential synergy between imaging- and gene-based hypoxia biomarkers, a better understanding of how their information relates to each other is crucial.

A major challenge in studies comparing the two methodologies is a shortage of paired imaging and gene data in patient cohorts; existing reports are few, based on small cohorts and do not address biopsy composition or intratumor heterogeneity (18). We have proposed imaging- and gene-based hypoxia biomarkers for cervical cancer patients, derived from DCE-MR images and gene expression data, respectively (19–22). The biomarkers have shown prognostic impact in several independent cohorts (19,21,23), and although they both inform about hypoxia, their underlying biology differs. The imaging biomarker depends on physiological features related to oxygen supply and consumption, such as blood perfusion, vascular density and cell density (24), while the gene-based biomarker measures expression of hypoxia responsive genes. In the present work, we utilized a unique paired data set of imaging- and gene-based biomarkers for 118 cervical cancer patients. We compared the performance of the two biomarkers in relation to the intratumor heterogeneity, and further investigated how they could be combined in prediction of treatment resistance.

## Materials and Methods

### Patient cohort

Totally 118 patients with locally advanced carcinomas of the uterine cervix, prospectively recruited to our chemoradiotherapy observational trial at the Norwegian Radium Hospital from 2001 to 2007, were included. The cohort constituted a representative subgroup of patients included in previous work to establish the gene-based biomarker (21), for which paired imaging and gene data were available (Supplementary Table S1). The study was approved by the Regional Committee for Medical and Health Research Ethics in southern Norway (S-01129). All patients gave written informed consent.

Treatment and follow-up were performed as described (20). In short, external radiation of 50 Gy in 25 fractions was given to the tumor, parametria and adjacent pelvic wall, while the remaining pelvis received 45 Gy. This was followed by brachytherapy of totally 25 Gy to the tumor in 5 fractions. Concurrent cisplatin (40 mg/m^2^) was given weekly in maximum 6 courses according to tolerance. Follow up was performed by standard procedures. When symptoms of relapse were noted, MRI of pelvis and retroperitoneum as well as X-ray of thorax were performed.

### Imaging-based biomarker

Diagnostic DCE-MR images for the imaging-based biomarker were acquired using a Signa Horizon LX-1.5T scanner (GE Medical Systems) with a pelvic-phased-array coil and a fast bolus injection of 0.1 mmol/kg body weight of Gd-DTPA (Magnevist®, Schering) (19,22). The dynamic T_1_-weighted series were acquired with a fast spoiled gradient recalled echo sequence and included 2–12 (median of 7) axial slices covering the whole tumor (Fig. 1A), with a slice thickness of 5 mm, slice gap of 1 mm, and in-plane resolution of 0.78 mm. Axial T_2_-weighted (T_2_W) images from a fast spin echo sequence were used for tumor delineation. The uptake of contrast agent; i.e. the relative signal intensity increase as a function of time after Gd-DTPA injection, was recorded for each voxel.

The A_Brix_-parameter in Brix pharmacokinetic model (24) was derived from the contrast uptake curves(19). Fraction of voxels in hypoxic regions was calculated from the A_Brix_-values of all tumor voxels, and this A_Brix_-hypoxic fraction was used as imaging biomarker (Fig. 1A). The A_Brix_ threshold value for hypoxia was identified in previous work by an iterative procedure to achieve the strongest correlation between hypoxic fraction and progression free survival (PFS) (22). By this procedure, all voxels with A_Brix_ below 1.56 were defined to locate in a hypoxic region (22), which could be visualized for each image slice in a binary A_Brix_-image (Fig. 1A). A cutoff for dichotomous classification of tumors was further identifed as the hypoxic fraction showing the strongest association to PFS. The cutoff was used to classify tumors according to their hypoxia status as more or less hypoxic (Fig. 1A).

**Figure 1.**
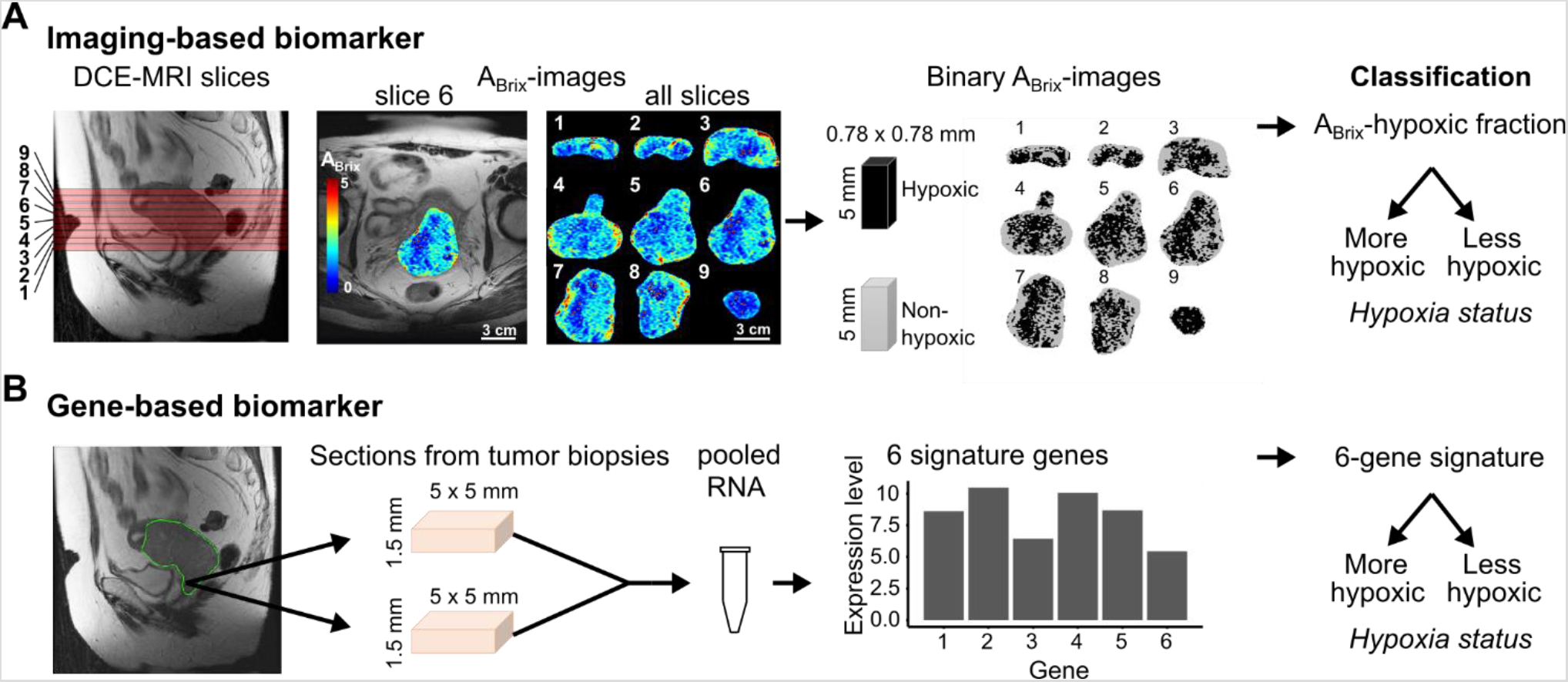
Imaging- and gene-based hypoxia biomarkers. **A**, Determination of the imaging-based biomarker from left to right, sagittal T2W-image of the pelvic showing localization of image slices numbered from lower to upper part of the tumor, example of axial ABrix-image of the tumor (slice 6) superimposed on T2W-image, ABrix-images of all slices covering the tumor, binary ABrix-images of the same slices showing voxels in hypoxic and non-hypoxic regions according to an ABrix threshold value of 1.56, and classification based on the hypoxic fraction of all slices combined. **B**, Determination of the gene-based biomarker from left to right, sagittal T2W-image of the pelvic showing localization of the region accessible for biopsies in the lower part of tumor, the approximately size of the sections from 1 – 4 biopsies (median 2) taken from each tumor and pooled for RNA isolation, expression data of 6 signature genes, and classification based on the signature value.

### Gene-based biomarker

Gene expression profiles for the gene-based biomarker were generated based on pooled RNA from 1–4 biopsies (median of 2) per tumor. The biopsies were taken from the lower, accessible region of the tumor the day after MRI (Fig. 1B), immediately snap frozen and stored at –80°C. From each biopsy, 30×50µm slices (approximately 5×5×1.5 mm) were used for RNA isolation. Assay methods for RNA isolation and gene expression measurement by Illumina beadarrays WG-6 v3 and HT-12 v4 (Illumina Inc.) were described previously (21). To compare classification from different biopsies within tumors, 2–4 biopsies from nine patients (24 biopsies in total) were available for individual analyses. Expression data were derived for each of these biopsies, using Illumina HT-12 v4 beadarrays and the same procedure as for the pooled samples. The data have been deposited in the gene expression omnibus (GEO) database (GSE146114). Tumor cell fraction, defined as the percentage of tumor cells in a haematoxylin and eosin stained section from the central part of the biopsy, was available for all biopsies.

The gene-based biomarker was constructed by evaluating the log_2_-transformed expression value of 31 hypoxia responsive genes associated with A_Brix_, and has been reported previously (21). In short, based on the ability of the expression values to separate patients correctly according to the A_Brix-_defined hypoxia status, a 6-gene signature (*DDIT3, ERO1A, KCTD11, P4HA2, STC2, UPK1A)* was identified and used as biomarker (21). The signature provides a continuous output value with a predefined cutoff of zero. This cutoff was used for dichotomous classification of tumors or samples according to their hypoxia status as more or less hypoxic (Fig. 1B).

### Heterogeneity and clustering of hypoxic regions

Intra- and inter-tumor heterogeneity in the biomarkers were estimated based on data for several samples per tumor, using random-effects one-way analysis-of-variance (ANOVA) models (25,26), where the total variance (T) is divided into the within-tumor (W) and between-tumor (B) variance. As a measure of the intratumor heterogeneity that is invariant to the measurement unit, W/T, where T=W+B, was used. Thus, a W/T>0.5 indicates a larger intratumor than intertumor heterogeneity, whereas for W/T<0.5 the heterogeneity within tumors is less than between tumors. When the biomarker value for each tumor is calculated based on the average of *k* samples instead of one, the within-tumor variance will be reduced by a factor 1/k, and thus the intratumor heterogeneity of the biomarker using these averages (W_k_/T) is given by:

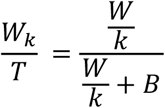

where W and B are the within- and between-tumor variance calculated using ANOVA with single sample values (26). ANOVA was performed on continuous biomarker values, and for the imaging-based biomarker, logit-transformed data were used.

Clustering of hypoxic regions was identified from the variance in imaging-based hypoxic fraction of numerous virtual samples randomly collected within an image slice. Each sample included 12 voxels, mimicking the size of a biopsy used for the gene-based biomarker. The number of virtual samples per patient was set to 1/12 of the total number of voxels in the corresponding slice. The variation in hypoxic fraction for the virtual samples from a tumor depends on the spatial distribution of the hypoxic ares as well as the hypoxic fraction of the slice, with lower variation for low and high hypoxic fractions. Therefore, to identify tumors with more and less clustering of hypoxic regions, the virtual sample variation adjusted for hypoxic fraction of whole slice was used as follows: The standard deviation (SD) of the hypoxic fractions for the virtual samples was calculated for each tumor and plotted versus the hypoxic fraction of the whole slice. Then, a generalized additive model (GAM) was fitted to the data to separate tumors with more and less clustering, defined as having positive and negative residuals, respectively.

### Composite biomarker score

To combine the imaging- and gene-based biomarkers into a composite hypoxia score for each patient, we used a dimension reduction approach. Based on a correlation plot of the continuous biomarker values, the optimal line for separation of the more and less hypoxic tumors to achieve the best association to PFS was determined:

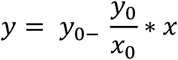

where *y* and *x* are the values of the two biomarkers and *y*_0_ and *x*_0_ are the points where the line intersects the *y*- and *x*-axis, respectively. The optimal separation line and, hence, *x*_0_ and *y*_0_, were determined in an iterative procedure, where the Cox proportional hazard (PH) model was used to test the difference in PFS between patients with more and less hypoxic tumors for numerous possible lines. A composite hypoxia score (*HS*_comp_) was found for each patient as the distance from the point (x,y) to the separation line:

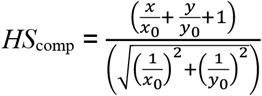

where less hypoxic tumors have *HS*_comp_ < 0 and more hypoxic tumors have *HS*_comp_ ≥ 0.

### Statistics

Clinical endpoint was PFS for follow-up until 5 years, where time from diagnosis to disease-related death or first event of relapse was used and patients were censored as described (21). Kaplan-Meier survival curves were compared using log-rank test. The univariate Cox PH model was used to determine hazard ratios (HR), and Cox uni- and multivariate PH analyses were performed to evaluate prognostic significance. Assumptions of PHs were confirmed graphically using log-minuslog plots.

Associations were estimated by Pearson’s or Spearman’s correlation and differences between groups were assessed with Fisher’s exact test, Wilcoxon rank-sum test or Kruskal-Wallis test, as appropriate. Significance level was 5%, and all tests were two-sided. Multivariate Cox PH analyses were performed in SPSS Statistics 21, while all other analyses were performed using R (27), version 3.6.0. HRs are presented with 95% confidence interval (CI).

## Results

### Relationship between imaging- and gene-based classification

Both the imaging- and gene-based biomarkers showed association to PFS when used as continuous variables in univariate Cox analysis (P=0.00024 and P=0.017, respectively). To classify patients into a less or more hypoxic group, a cutoff hypoxic fraction of 0.27 was used for the imaging-based biomarker, leading to the strongest association to outcome (Supplementary Fig. S1). The predefined cutoff of zero (21) was used for the gene-based biomarker. Classification with these thresholds yielded 36 (31%) and 43 (36%) patients in the more hypoxic group based on imaging and genes, respectively. Thus, the gene-based biomarker classified a higher number of tumors as more hypoxic. Both biomarkers revealed a highly significant difference in outcome between the patient groups (Fig. 2A, B).

**Figure 2.**
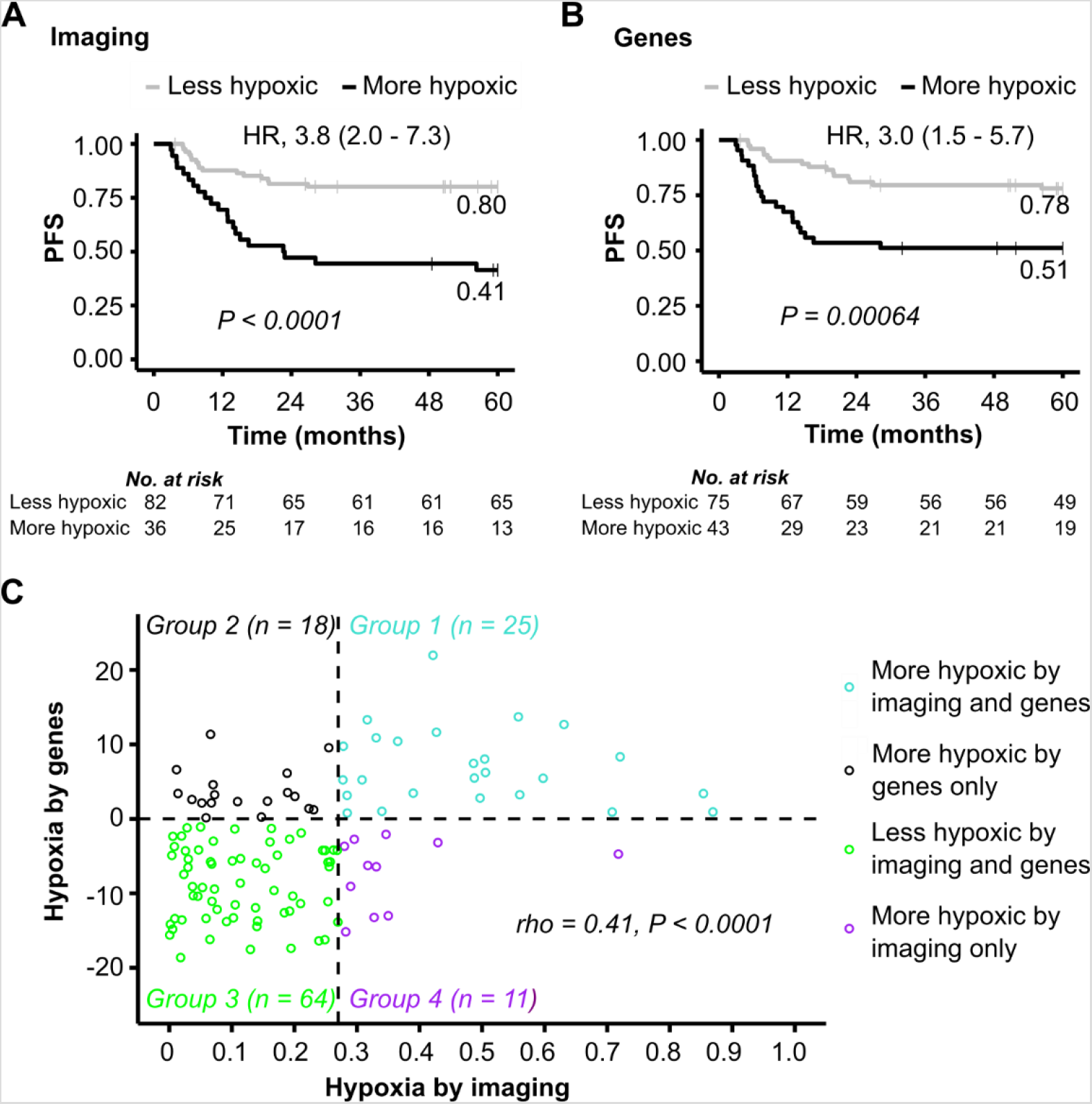
Comparison of imaging-and gene-based hypoxia classification. Progression free survival (PFS) for 118 patients with more or less hypoxic tumors based on imaging (**A**) and genes (**B**). *P*-values from log-rank tests, HR with 95% CI from Cox PH analysis and number of patients at risk are indicated. **C**, Correlation plot of biomarker values, showing gene-defined *versus* imaging-defined hypoxia for 118 patients. Dotted lines, classification cutoff of each biomarker, defining four groups according to the similarity in classification by imaging and genes. *P*-value and regression coefficient (*rho*) from Spearman correlation analysis are indicated.

A correlation plot of gene- *versus* imaging-defined hypoxia was generated to compare classification by the two biomarkers for individual patients. A significant correlation was found (Fig. 2C), and this correlation remained significant after removing 41 tumors used in previous work to construct the gene-based biomarker (21) (Supplementary Fig. S2). Four classification groups were defined; group 1 tumors were more hypoxic by both biomarkers, group 2 tumors were more hypoxic by genes, but less hypoxic by imaging, group 3 tumors were less hypoxic by both biomarkers, and group 4 tumors were less hypoxic by genes, but more hypoxic by imaging (Fig. 2C). There was a significant association between the classifications by the two biomarkers (*P*<0.0001; Fisher’s exact test), and for 89 of the 118 tumors (75%), imaging and genes classified the tumors into the same group; 25 (21%) as more hypoxic (group 1) and 64 (54%) as less hypoxic (group 3). Inconsistent classification (29 tumors; groups 2, 4) tended to be more frequent for the imaging-defined more hypoxic tumors than the less hypoxic tumors (31% versus 22%), but this difference was not statistically significant (Fisher’s exact test). Hence, although equal classification by the two biomarkers was generally achieved, a group of patients was differently classified regardless of the hypoxia status of their tumor.

### Heterogeneity in hypoxia across the tumor volume

Inconsistent classification by the two biomarkers could be more common in large than small tumors, since the biopsies include only a small part of the tumor, and may therefore have lower ability to record the hypoxia status correctly in these cases. In contrast to this hypothesis, the tumors classified as more hypoxic by both biomarkers (group 1) had a larger volume than the others, and there was no significant difference in the volume between groups 2–4 (Fig. 3A). A detailed investigation of regional differences in hypoxia was therefore performed by first using the imaging data to assess the heterogeneity across image slices. The binary images revealed pronounced differences up to 0.54 in hypoxic fraction between slices of individual tumors, and the hypoxia status of each slice could differ (Fig. 3B-C; Supplementary Fig. S3). However, hypoxic fraction per slice showed a small intratumor heterogeneity with a W/T of 0.16. Moreover, most slices (88%) were classified with the same hypoxia status as the whole tumor (Fig. 3C).

**Figure 3.**
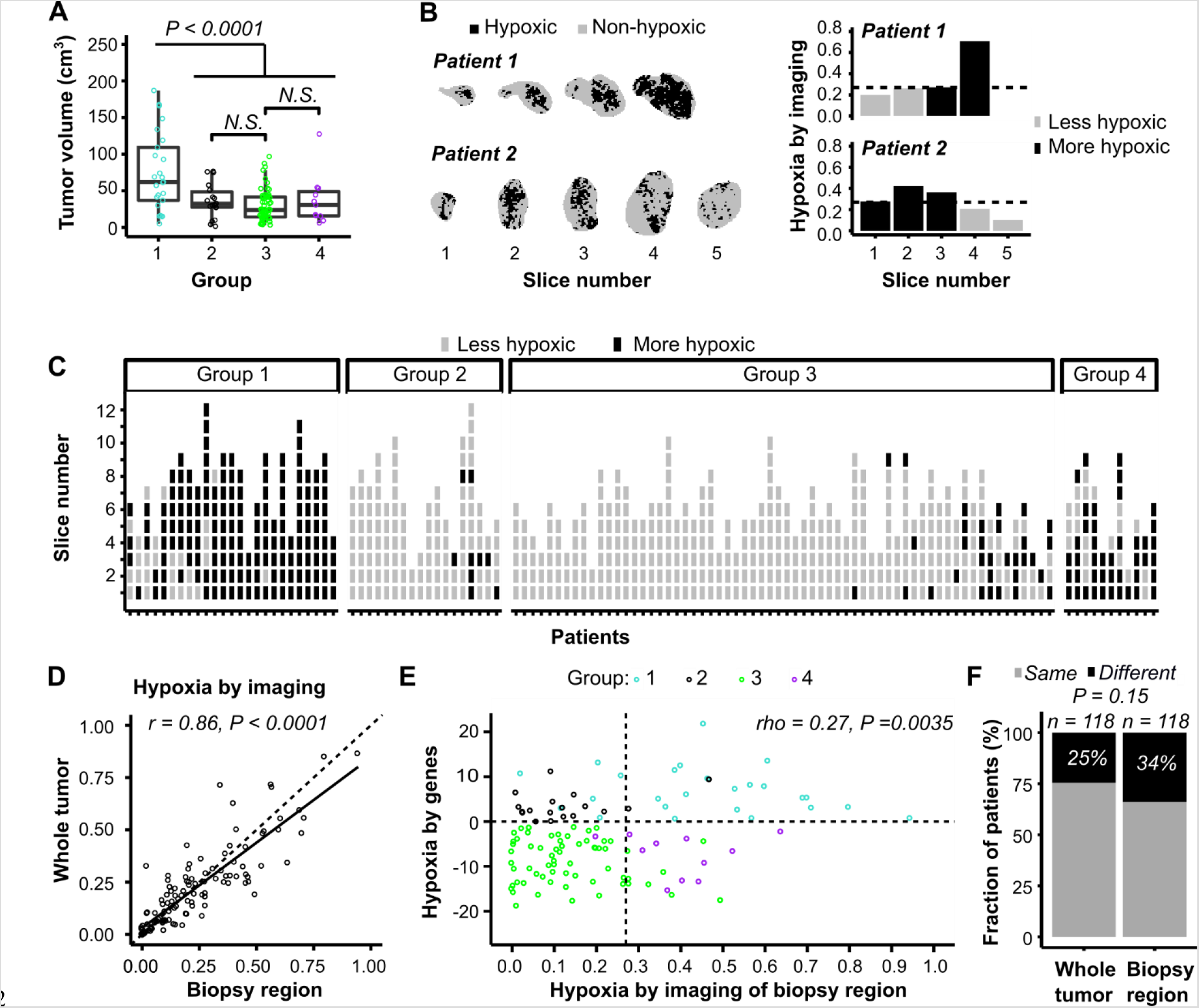
Heterogeneity in hypoxia across image slices. **A**, Tumor volume for the four classification groups defined in Fig. 2C. Jitterplot overlaid a boxplot, where the boxes extend from the first to third quartile with the median value indicated. *P*-value from Wilcoxon rank-sum test is indicated. N.S., non-significant. **B**, Binary images showing voxels in hypoxic and non-hypoxic regions of two patients with heterogeneous hypoxia status across image slices (left), and imaging-defined hypoxia (hypoxic fraction) of each slice (right). Dotted lines, classification cutoff (0.27). **C**, Classification of individual image slices by the imaging-based biomarker. The patients are devided into four classification groups defined in Fig. 2C, and are sorted according to increasing biomarker value of the whole tumor for each group. **D**, Correlation between imaging defined hypoxia of the biopsy region and the whole tumor. Dotted line, 1:1 relationship; solid line, linear regression line. **E**, Comparison of the imaging- and gene-based biomarker values using image slice 1 from the biopsy region. Dotted lines, classification cutoff of each biomarker. The four classification groups defined in Fig. 2C are indicated by colors. **F**, Fraction of patients classified with same or different hypoxia status by imaging and genes, using images from the whole tumor or the biopsy region. Number of patients (*n*) and P-value from Fisher’s exact test are indicated. **D, E**, *P*-value and regression coefficient from Pearson (**D**) and Spearman (**E**) correlation analysis are indicated.

The image slice covering the biopsy region (i.e. slice 1) was selected to investigate how well the hypoxia status of this region reflected the status of the whole tumor. Imaging-defined hypoxia based on this slice showed a strong correlation with the value based on all slices (r = 0.86, P<0.0001; Fig. 3D). Moreover, both a significant correlation between imaging- and gene-defined hypoxia (rho = 0.27, P = 0.0035; Fig. 3E) as well as the similarity in classification by imaging and genes (Fig. 3F) were retained, but not improved, when the imaging data were derived from the biopsy region rather than the whole tumor. The hypoxia status of the biopsy region therefore seemed to be representative for the whole tumor. Moreover, the gene-based biomarker reflected equally well the imaging-defined hypoxia status of the whole tumor and the biopsy region.

### Heterogeneity in hypoxia within the biopsy region

The heterogeneity in hypoxia was further investigated within the biopsy region. In a subgroup of nine patients, hypoxia was assessed by genes for multiple biopsies from each tumor (Fig. 4A). The intratumor heterogeneity in gene-defined hypoxia was smaller than the heterogeneity between tumors with a W/T of 0.33. By using 2–4 biopsies, W/T was reduced to 0.20 (2 biopsies), 0.14 (3 biopsies) and 0.11 (4 biopsies). However, the number of biopsies used for patient classification in Figure 2 (*i.e*., 1–4) was not associated with the similarity in imaging- and gene-based classification (Supplementary Fig. S4). Moreover, for eight of nine tumors in our multiple biopsy experiment there was a complete concordance in classification among the biopsies (Fig. 4B). For the remaining tumor (no. 8, Fig. 4B), one biopsy was classified as more hypoxic and two as less hypoxic. The binary image of the slice covering the biopsy region (slice 1) of this tumor showed large spatial variation with clustering of hypoxic areas (Fig. 4C), which could imply that the biomarker value strongly depended on the exact location of the biopsy.

**Figure 4.**
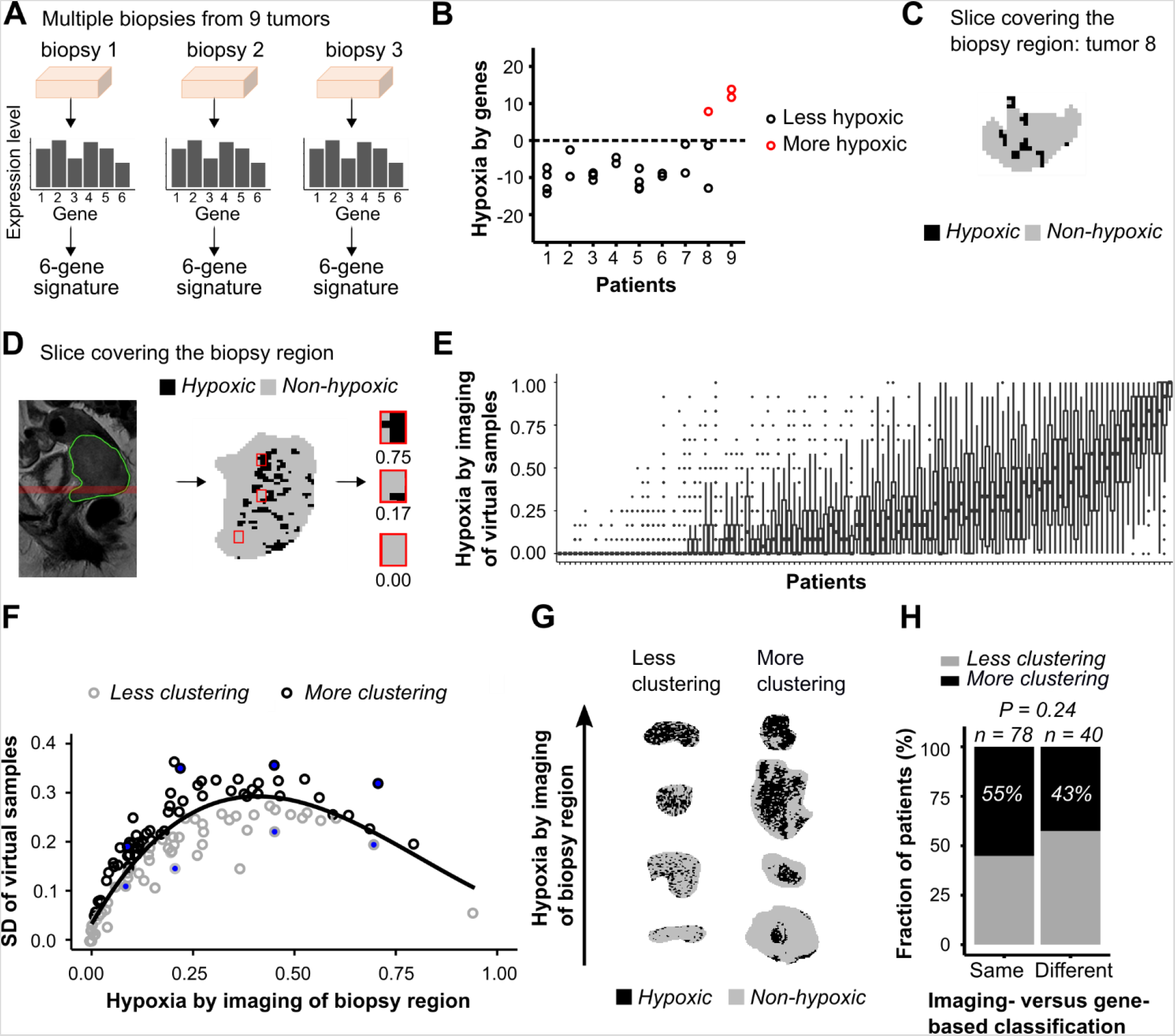
Heterogeneity in hypoxia within biopsy region. **A**, Determination of gene-based biomarker for multiple biopsies from 9 tumors. **B**, Gene-based biomarker value for 2–4 biopsies from each of 9 tumors (24 samples). Dotted line, classification cutoff. **C**, Binary image of the biopsy region for tumor 8 in (**B**). **D**, Illustration of the simulation experiment. Sagittal T2W-image of the pelvic with indication of selected image slice covering the biopsy region (left), binary image of selected slice showing voxels in hypoxic and non-hypoxic regions and location of 3 virtual samples, each of 12 voxels (middle), binary image and biomarker value (hypoxic fraction) of the 3 virtual samples (right). **E**, Imaging-based biomarker value of the virtual samples for each patient sorted according to increasing biomarker value of the biopsy region. The boxes extend from the first to third quartile with the median value indicated. **F**, Standard deviation (SD) of the imaging-based biomarker value of the virtual samples *versus* biomarker value of the biopsy region. Line, generalized additive model (GAM) fitted to the data to separate tumors with more (above the line) or less (below the line) clustering of hypoxic regions. Filled circles, tumors displayed in (**G**). **G**, Binary images showing less or more clustering of hypoxic regions for 8 tumors indicated in (**F**). **H**, Fraction of patients with less or more clustering of hypoxic regions for patients with same or different hypoxia status by imaging (biopsy region) and genes. Number of patients (*n*) and P-value from Fisher’s exact test are indicated.

To address this hypothesis, we performed a simulation experiment to assess the spatial variation in hypoxia within the biopsy region using the imaging data of slice 1. We randomly sampled numerous areas of biopsy size, each of 12 voxels, within the slice and determined the hypoxic fraction for each of these virtual samples (Fig. 4D-E). The virtual samples showed a large intratumor heterogeneity, with a W/T of 0.50, indicating high spatial variation in imaging-defined hypoxia across the biopsy region. Tumors with less or more clustering of hypoxic regions were identified by comparing the variation in the imaging-defined hypoxia of the virtual samples for each tumor, correcting for the whole-slice hypoxic fraction (Fig. 4F). Considerable difference in the degree of clustering was found across the tumors, in agreement with a visual inspection of the image slices (Fig. 4G; Supplementary Fig. S5-S6). Clustering was, however, not associated with inconsistent classification by imaging and genes (Fig. 4H). Overall, the gene-based biomarker seemed to be reproducible across biopsies, and spatial variation in hypoxia within the biopsy region could not explain differences in classification by the two biomarkers.

### Heterogeneity in biopsy composition and gene-based hypoxia

Tumor cell fraction was 50% or higher in most biopsies underlying the gene-based biomarker, however, it showed a broad range from 10% to 90% (median of 65%). No correlation between this fraction and gene-defined hypoxia was found (Fig. 5A). In particular, both more and less hypoxic tumors were identified at tumor cell fractions up to 90% and down to 40%. All three tumors with a fraction below 40% were classified as less hypoxic by genes, but these were also less hypoxic by imaging (group 3; Fig. 5A). Moreover, there was no significant difference in this fraction between any of the classification groups (Fig. 5B). A more detailed analysis was also performed, using data from our multiple biopsy experiment (Fig. 4A). Although the fraction could differ up to 40% for some biopsies from the same tumor (Fig. 5C, tumors no. 3 and 8), they were generally classified with the same hypoxia status. For tumor no. 8, the only biopsy out of three that was classified as more hypoxic had the lowest tumor cell fraction of 20%. Altogether, the gene-based classification seemed to be independent on biopsy composition.

**Figure 5.**
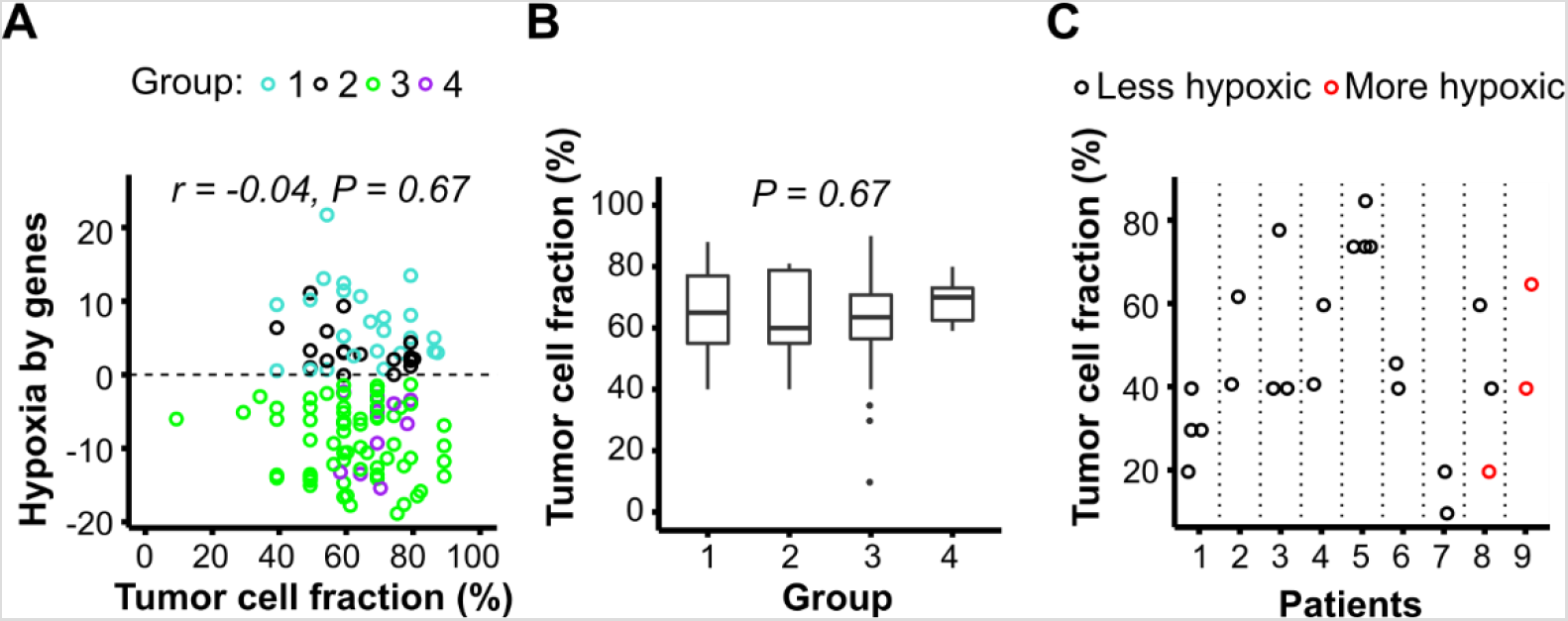
Tumor cell fraction and gene-based classification. **A**, Gene-based biomarker value *versus* tumor cell fraction in biopsy (n = 118). Mean fraction of multiple biopsies used for classification of the tumor is shown. The color indicates the four classification groups defined in Fig. 2C. Dotted line, classification cutoff. *P*-value and regression coefficient (*r*) from Pearson correlation analysis are indicated. **B**, Tumor cell fraction of the four classification groups. The boxes extend from the first to third quartile with the median value indicated. P-value from Kruskal-Wallis test is indicated. **C**, Tumor cell fraction and hypoxia status by the gene-based biomarker of individual biopsies from 9 tumors presented in Fig. 4B.

### Patient classification based on a combined biomarker

The above results indicated considerable robustness of the two biomarkers, where the different classification of some tumors seemed not to be caused by intratumor heterogeneity. It is therefore likely that the biomarkers provide complementary information that combined could lead to more precise prediction of treatment outcome. To test this hypothesis, we first compared the survival curves for the four classification groups (Fig. 6A). Patients with a more hypoxic tumor by both imaging and genes (group 1) showed poor survival compared to the others with a 60 months PFS of 0.32. Moreover, for patients with a group 2 or 4 tumor, which was classified as more hypoxic by one biomarker only, the outcome was much better with a 60 months PFS of 0.78 and 0.64, respectively. This outcome was close to the PFS of 0.81 for patients with a less hypoxic tumor by both biomarkers (group 3). Combination of the two biomarkers by requiring more hypoxia by both imaging and genes to define the more hypoxic tumors, i.e. comparing group 1 with group 2–4, improved the prognostic impact from a HR of 3.8 (imaging) and 3.0 (genes) to a HR of 4.8 (Fig. 6B).

**Figure 6.**
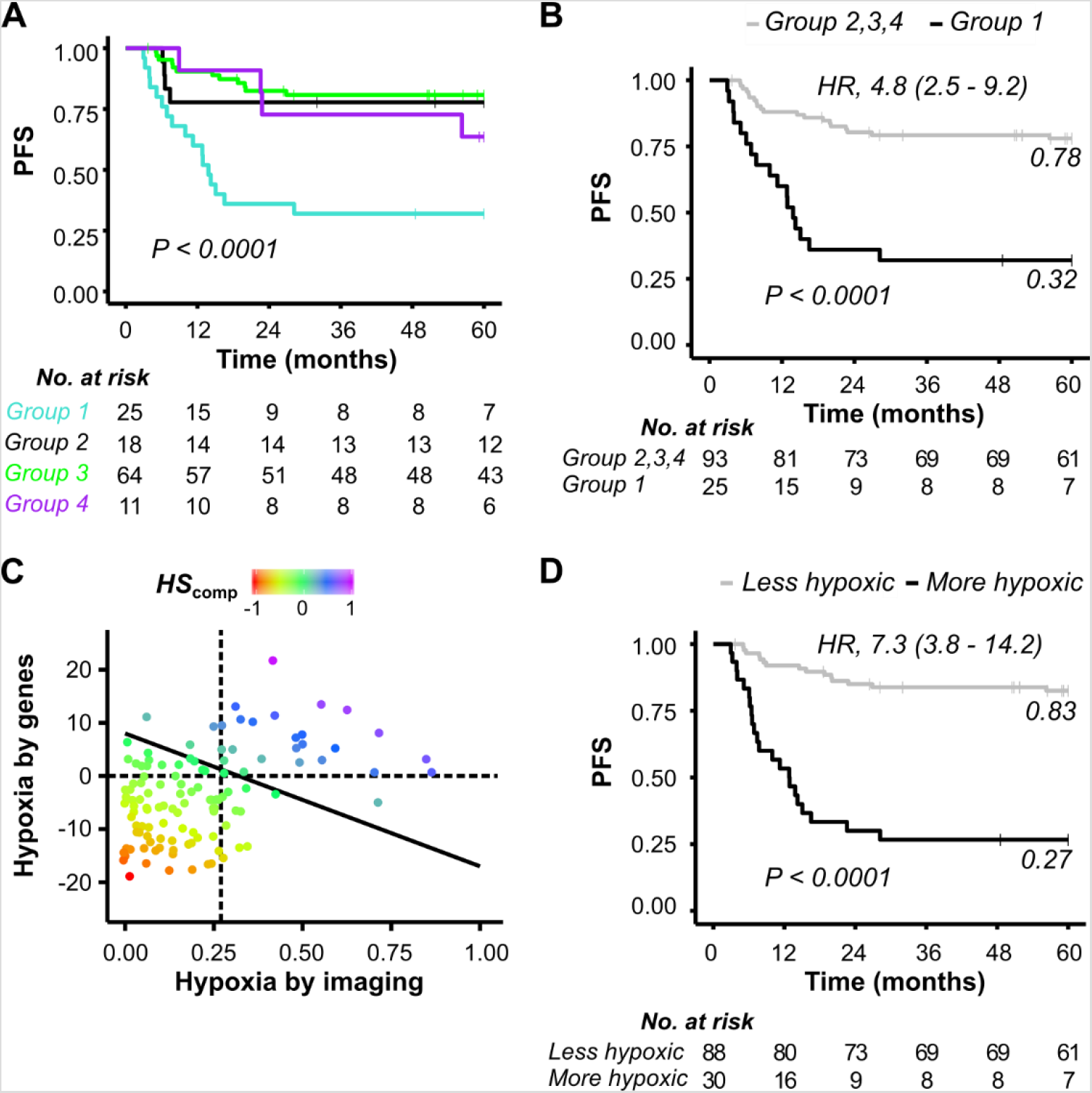
Combined imaging- and gene-based biomarker. **A**, Kaplan-Meier curves for progression-free survival (PFS) of patients in the four classification groups defined in Fig. 2C. **B**, Kaplan-Meier curves for PFS of patients classified with more hypoxic tumor by both imaging and genes (group 1), and with less hypoxic tumor by at least one biomarker (group 2,3,4). **C**, Correlation plot of biomarker values, showing gene-defined *versus* imaging-defined hypoxia. The optimal line for classifying patients with a more (above the line) and less (below the line) hypoxic tumor to achieve the strongest association to PFS is shown. **D**, Kaplan-Meier curves for PFS of patients classified into the two groups defined in (**C**). **A, B, D**, *P*-values from log-rank test and number of patients at risk are indicated. **B, D**, 60-month recurrence probability and HR with 95% CIs are indicated.

Some patients with more hypoxic tumor by only one biomarker had treatment failure. To investigate the possibility to identify these cases as high risk patients, we utilized a dimension reduction approach to combine the two biomarkers. In a correlation plot of the biomarker values, the line separating the more and less hypoxic tumors to yield the strongest association to PFS was found (Fig. 6C; Supplementary Fig. S7). With this separation, six tumors originally defined as more hypoxic by one biomarker only (group 2 and 4) were classified to the more hypoxic group. The other groups remained unchanged, except that one tumor with borderline more hypoxia by both biomarkers moved to the less hypoxic group. The resulting continuous composite hypoxia score (*HS*_comp_) was prognostic (P = 0.00075), i.e. there was an increased risk of recurrence with increasing score. Hypoxia status defined from *HS*_comp_ gave a difference in PFS at 60 months of 0.56 between patients with more or less hypoxic tumor (P<0.0001, HR=7.3) and survival probabilities of 0.27 and 0.83 for these patients, respectively (Fig. 6D). Our new strategy to combine the two biomarkers therefore further increased the prognostic impact of hypoxia classification. Moreover, the hypoxia status defined from *HS*_comp_ showed a strong prognostic value in multivariate analyses together with the clinical parameters FIGO stage, lymph node involvement, and tumor volume (Supplementary Table S2).

## Discussion

The present study is the first to report combination of an imaging- and a gene-based biomarker for classification of cancer patients according to a specific tumor phenotype, and further to address the influence of intratumor heterogeneity. Although of totally different origin, imaging- and gene-based biomarkers are both used in clinical trials to identify patients with hypoxic tumor, emphasizing the need for a better understanding of how they relate to each other. Access to a large, paired data set on previously validated hypoxia biomarkers in our study enabled reliable comparison of the two biomarkers and evaluation of their prognostic potential in a combined setting. Moreover, by utilizing an imaging biomarker constructed from MR images with high spatial resolution compared to biopsy size, intratumor heterogeneity in hypoxia could be assessed on a scale of relevance for the gene-based biomarker. Although considerable heterogeneity was found, this seemed to have no major influence on the performance of the two biomarkers, and a synergy in prediction of hypoxia-related treatment resistance was demonstrated.

The imaging data suggested large regional differences in hypoxia across the tumor volume. This is consistent with oxygen tension (pO_2_) measurements by electrodes, showing broad pO_2_ distributions of individual cervix tumors (28–30). Considerable heterogeneity in pO_2_ has also been reported for sarcomas and cancer of the head and neck and breast (31–33), and regional differences in hypoxia is a common feature of solid tumors (34). Although this has caused concern for the application of hypoxia biomarkers derived from only a limited part of the tumor (35), our study showed low heterogeneity in hypoxic fraction across all image slices, with a W/T of 0.16, as well as good agreement between the hypoxia status of the biopsy region and the whole tumor. Moreover, consistent classification by imaging and genes was found for almost all of the largest tumors, showing representative hypoxia status of the biopsy also in these cases. In accordance with our results, no significant difference was found in a study comparing pO_2_ data from three different depths in cervix tumors (36). It therefore seems to be a minor hurdle for application of the gene- based biomarker that only the lower part of the tumor is accessible for biopsies and the tumors are large compared to biopsy size.

Imaging data of the virtual samples further revealed considerable heterogeneity in hypoxia within the biopsy region, consistent with reports of a large variation in pO_2_ along electrode tracks of 5–10 mm in cervical and head and neck tumors (31,37). Imaging has been used to quantify intratumor heterogeneity parameters, like clustering of voxel-wise data, by several approaches and with different degree of complexity (13). In previous work we investigated imaging-defined heterogeneity in cervical cancer, but the hypoxic fraction of each tumor was not taken into account (38,39). In the present work we assessed clustering of hypoxic regions at the biopsy level by analyzing the imaging-based biomarker and its variance for image-areas of biopsy size, since this approach provided a direct link to our gene-based biomarker. Moreover, since the variance was dependent on hypoxic fraction, this was taken into account when identifying tumors with more and less clustering. Severe clustering was indicated for many tumors, but appeared to play no major role for an inconsistent classification by imaging and genes. The same conclusion could be drawn regarding a heterogeneity in tumor cell fraction across biopsies. Hence, no relationship to the gene-based biomarker was found, in accordance with a previous report on hypoxia classification by gene expression in head and neck cancer (40). However, it should be emphasized that heterogeneity in hypoxia or biopsy composition may have led to different classification by the two biomarkers in a few cases.

The gene-based biomarker showed a low intratumor heterogeneity, with an estimated W/T of 0.33 for a single biopsy. This is consistent with reports showing that multigene signatures tend to have lower heterogeneity than individual genes (41). When using up to 4 biopsies, W/T was reduced to around the 0.15 limit suggested for a biomarker to represent the tumor with satisfactory accuracy(42). However, the W/T is based on the continuous biomarker values. For hypoxia classification, the biomarker seems to be robust also when based on a single biopsy. The use of 3–4 rather than 1 or 2 biopsies was not associated with more consistent classification with imaging and genes. Moreover, for 8 of 9 patients in our multiple biopsy experiment, all biopsies from the same tumor were classified with the same hypoxia status. It therefore seems that our gene-based biomarker captured a molecular hypoxia phenotype characteristic of the tumor, independent of the number of biopsies used.

The imaging- and gene-based biomarkers assess hypoxia at the physiological and molecular level, respectively. By imaging, a surrogate of hypoxia is recorded as a balance between oxygen consumption and supply parameters regardless of molecular features. In contrast, expression of hypoxia responsive genes, as measured by the gene-based biomarker, is influenced by persistent genetic alterations such as DNA methylation, mutations and copy number changes, in addition to an instant stimulatory effect on transcription by reduced oxygen concentration. One could therefore speculate that the gene-based biomarker reflects a phenotype providing tolerance of tumor cells to hypoxic stress, and thereby capability to tackle fluctuations in hypoxia. By such hypothesis, the highly aggressive tumors would be those with both low oxygen concentration, as recorded at the time of imaging, and high hypoxia tolerance. Our outcome data for the combined biomarker support this hypothesis, showing the worse survival of patients classified with hypoxia by both biomarkers.

The combined biomarker improved risk classification of the patients, probably by reflecting different hypoxia phenotypes related to tumor aggressiveness. In addition, combining the biomarkers likely reduced small contributions from intratumor heterogeneity and uncertainties caused by technical factors and changes in hypoxia status during the time period between MRI and collection of biopsies (13,26). One way to combine two biomarkers is to require equal classification by both of them to conclude about risk status, which was a successful strategy in our study. In addition, we presented an alternative approach by using dimension reduction to calculate a composite score, where patients with a highly hypoxic tumor by only one of the biomarkers were moved to the more hypoxic group. This strategy led to the strongest association to outcome and could therefore be of particular interest for the clinical implementation of the biomarkers. Moreover, the risk of failure increased with increasing score, showing that the score could be of value for biological analyses of tumors in addition to classification of patients. The approach may also be exploited for other phenotypes than hypoxia, and can possibly help development of robust biomarkers for personalized medicine (4).

Tumor hypoxia is a major factor associated with treatment resistance and metastasis in cervical cancer (43–47) and therefore an attractive target for intervention. Several hypoxia targeting drugs have reached clinical trials (10,48), but the results so far have been disappointing. Individual genes in our gene-based biomarker inform about key pathways in the hypoxia response of cervix tumors, including the hypoxia inducible factor HIF1A pathway and the unfolded protein response (UPR), for which promising targeting drugs are in pipeline (21). Such information could indicate the best pathway to target and thereby complement the imaging data. This might be of utmost importance since cervical cancer patients are already on the toxicity limit with the standard treatment and should not enter clinical trials without an expected benefit (49,50). Our combined biomarker may therefore lead to a better trial design, both by providing more accurate identification of patients with treatment resistant disease and by proposing the most relevant hypoxia targeting drugs for intervention.

## Data Availability

The data have been deposited in the gene expression omnibus (GEO) database (GSE146114), which will be released upon publication.

